# Ultra-processed food consumption and frailty in older adults: a systematic review and meta-analysis

**DOI:** 10.64898/2026.03.29.26349639

**Authors:** Min Pu, Qianying Ma

## Abstract

Frailty is an age-related syndrome characterized by biological dysfunction and reduced physiological reserve in response to stressors. Its prevalence is increasing with population aging, resulting in a substantial health burden due to adverse outcomes on health, such as cardiovascular disease and mortality. Ultra-processed foods (UPFs), defined as industrial formulations made primarily from processed ingredients, have received increasing attention due to their potential role in the development and progression of frailty. This systematic review and meta-analysis examined the association between ultra-processed food intake and the risk of frailty in older adults. This study systematically searched for all relevant studies published up to January 2026. Ten observational studies involving 105327 participants, comprising 6 prospective and 4 cross-sectional studies, were included in the systematic review, of which 6 were eligible for meta-analysis. Random-effects models were employed to estimate pooled effect sizes and 95% confidence intervals (95% CIs). Meta-analysis showed that higher consumption of UPFs was significantly associated with an increased risk of frailty (pooled OR = 1.43, 95% CI = [1.02-2.005], p = 0.041). Narrative synthesis further supported a positive association between UPF intake and frailty or related outcomes. Our findings suggest that a higher consumption of ultra-processed foods may contribute to frailty risk, potentially through inflammatory pathways. However, given the high heterogeneity, results should be interpreted with caution. Overall, our findings suggest that reducing UPF consumption may be a promising target for public health strategies to prevent frailty in ageing populations.

## Introduction

Globally, frailty affects approximately 12% of older adults (Caoimh et al., 2021), and its prevalence is expected to rise as the global population ages rapidly. Frailty is characterized by reduced biological and physiological functioning and increased vulnerability to stress (Chen et al., 2014). It is commonly defined by the Fried frailty phenotype, which comprises multiple symptoms including limited physical activity, weakness, exhaustion, slow walking speed, and unintentional weight loss (Fried et al., 2001). Individuals testing positive with three or more of these symptoms are classified as frail. Growing evidence shows that frailty is associated with adverse health outcomes, such as increased risks of hospitalization, death, and mental disorders (Jiang et al., 2024), leading to substantial health burden and health-care costs. Therefore, understanding the factors underlying frailty at the population level is critical.

Among modifiable factors influencing frailty, dietary intake is particularly important because adequate nutrition is essential for maintaining physiological homeostasis. However, modern dietary patterns rely excessively on ultra-processed foods (UPFs). The UPFs are strongly associated with poor diet quality, as they are typically high in added sugars, saturated fats, and sodium, and low in essential nutrients commonly found in whole foods (Monteiro et al., 2019). The NOVA classification system is a framework for categorizing foods according to the extent and purpose of their processing. Within this framework, UPFs are defined as hyperpalatable formulations produced through industrial processes (Monteiro et al., 2019). Despite poor nutritional profiles, UPFs are increasingly prevalent in modern diets, largely due to their convenience (e.g., being ready-to-eat), low cost, and hyperpalatability. The overconsumption of ultra-processed foods has been associated with adverse health outcomes in older adults (Mariath et al., 2022). UPF intake has been linked to low-grade systemic inflammation (Malesza et al., 2021) and an increased risk of chronic diseases such as obesity and type 2 diabetes (Gutierrez-Ortiz et al., 2025; Lane et al., 2024), cardiovascular diseases (Souza et al., 2022), hypertension (Wang et al., 2022), and mental health disorders (Mengist et al., 2025).

Notably, emerging empirical evidence suggests that ultra-processed foods are closely associated with frailty. An early prospective study found that higher consumption of UPFs, as quantified by energy intake and grams per day/body weight, was a strong predictor of frailty risk over a 3.5-year follow-up period in Spain (Sandoval-Insausti, Jiménez-Onsurbe, et al., 2020), suggesting a potential causal relationship between UPFs and frailty development. Subsequent studies in the United States populations have similarly revealed a positive association between UPFs and frailty. For instance, a cross-sectional study found that a 10% increase in energy intake from UPFs was associated with a 0.02-fold increase in the risk of pre-frailty or frailty among non-obese participants (Hao et al., 2022). In contrast, another study found no significant association between UPF consumption and frailty, although higher intake of unprocessed or minimally processed foods was associated with a lower risk of frailty (Zupo et al., 2023). These inconsistent findings highlight the need to systematically incorporate relevant studies to better understand the effect of UPFs on frailty.

To our knowledge, evidence synthesizing the association between UPF consumption and frailty remains limited, and a comprehensive quantitative synthesis of this relationship is lacking. Therefore, it is imperative to objectively evaluate the association through a systematic review and meta-analysis. Addressing this gap, this study aims to assess and quantify the relationship between UPFs and frailty prevalence. The findings may inform targeted interventions focusing on modifiable dietary factors, such as reducing UPF intake, to prevent or potentially reverse frailty in older adults (Lochlainn et al., 2021).

## Methods

### Search strategy

This review was conducted following the Preferred Items for Systematic Reviews and Meta-Analysis (PRISMA) guideline (Page et al., 2021). PubMed, Web of Science, and Google Scholar were searched for articles published up to 5 January 2026. Google Scholar was searched as a supplementary source to identify additional relevant studies. The search term used was (“ultra-processed food*” OR “ultraprocessed food*” OR UPF OR “NOVA” OR “NOVA classification” OR “NOVA group 4” OR “processed food*” OR “highly processed food*”) AND (frailty OR frail OR “frailty index” OR “frailty phenotype” OR “physical frailty”). To further identify potentially missed publications, we also reviewed relevant references listed by all selected studies. The details of the literature search are shown in **Figure 1**.

**Figure 1.**
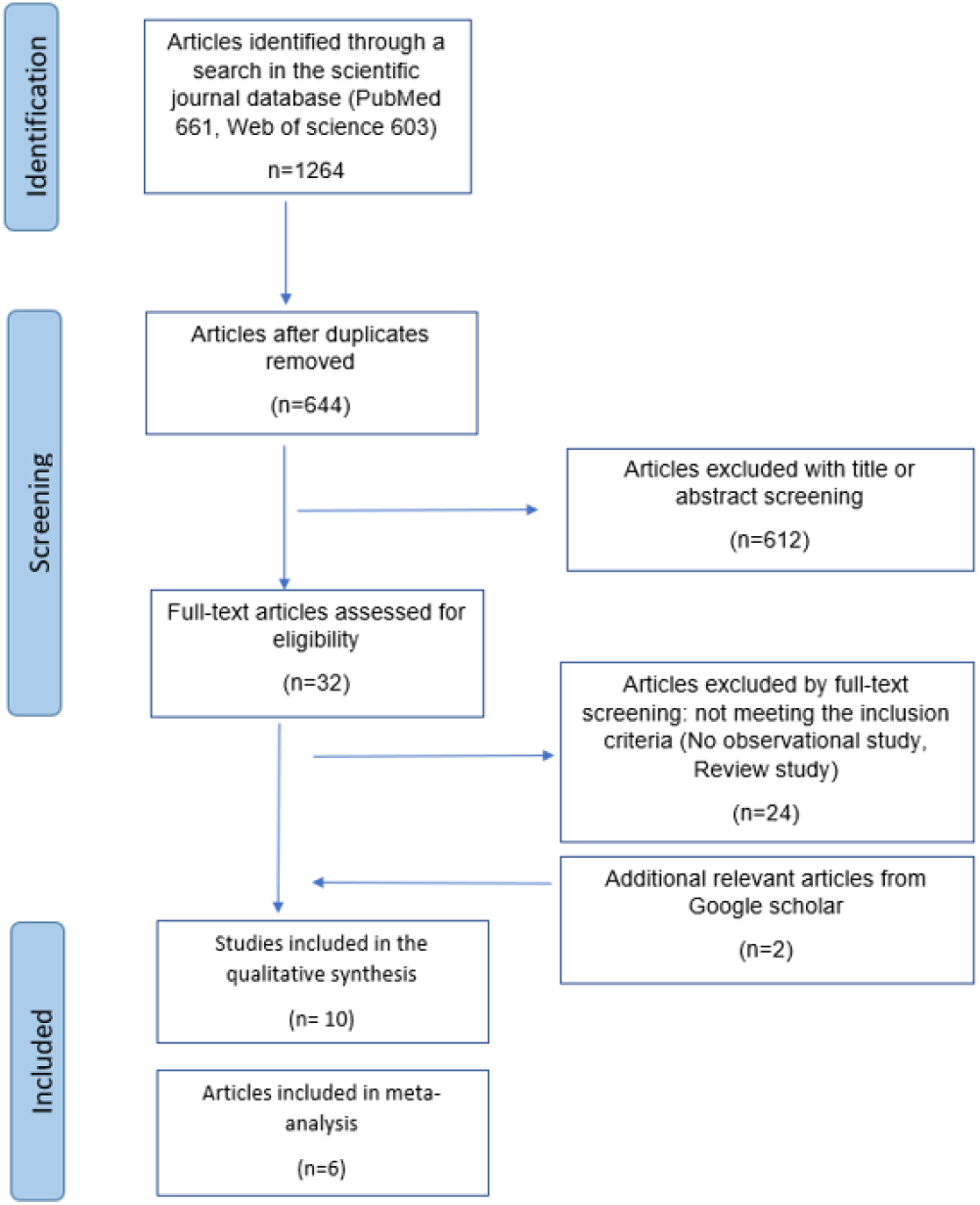
Literature search flow chart.

### Inclusion and exclusion criteria

The following inclusion criteria were implemented: (1) observational studies; (2) publications in English; (3) studies considering ultra-processed food as the exposure, as assessed by dietary intake assessment and the NOVA classification systems; (4) studies investigating the impact of ultra-processed foods on frailty; (5) studies conducted on older adults; (6) studies providing sufficient data to compute effect sizes, such as reporting odds ratio (OR), relative risks (RRs), hazard ratio (HR), or β, together with corresponding 95% confidence intervals (CIs). We also implemented the following exclusion criteria: (1) studies conducted on clinical human samples; (2) studies that reported insufficient data for computing effect size; (3) review articles, comments, letters; (4) non-observational study; (5) review study; (6) non-English study.

### Data extraction

We extracted the following variables based a structured data extraction form: (1) Study characteristics including authors, year of publication; (2) study characteristics including sample size, study location, age, follow-up duration, and sex; (3) study design (cross-sectional, prospective); (4) assessment methods for ultra-processed food exposure; (5) assessment methods for frailty; (6) the results assessing UPF intake and risk of frailty, including the ORs, HR, RRs, and 95% confidence intervals. Reported risk estimates for frailty with the highest exposure to UPFs were compared with those with the lowest exposure levels to UPFs (as assessed by the NOVA classification system), while adjusted estimates of covariates were extracted.

### Quality of evidence

We applied the Newcastle-Ottawa Scale (NOS) to assess the overall quality of studies. The NOS checklist includes three sections: selection, comparability, and outcome, with a maximum of four, two, and three points, respectively. Poor quality is 1-3 points, fair quality is 4-6 points, and high quality is 7-9 points (Stang, 2010).

### Statistical analysis

Six publications reported OR and 95% CI were included in the meta-analysis, while an additional four papers reporting RR, HR, or β were included in the narrative synthesis. In the meta-analysis, all data were expressed as log-odds ratios (log ORs) with standard errors (SEs). For studies reporting multiple effect sizes based on different measures of UPFs, we included UPF energy intake percentage for the meta-analysis, as this was the most commonly reported measure across the included studies. A random-effects model was used to pool effect sizes and corresponding 95% CIs when significant heterogeneity was present. The potential heterogeneity among studies was assessed using the Q test and I^2^ . Given the small number of included studies, we did not conduct subgroup meta-analysis. All analyses were conducted in R Studio using the rma.mv function of the “metafor” R package (Viechtbauer, 2010).

## Results

### Study characteristic

A total of 10 studies were included in this systematic review, comprising 105372 participants. Information on the included studies is presented in **Table 1**. Three studies were conducted in the United States (Fung et al., 2024; Hao et al., 2022; Konieczynski et al., 2025), while the remaining studies were conducted in Spain (Sandoval-Insausti, Blanco-Rojo, et al., 2020), Italy (X. Li et al., 2025; Zupo et al., 2023), Australia (Clayton-Chubb et al., 2024), Singapore (Y. Li et al., 2025), Iran (Hallajzadeh et al., 2025), and China (Zhang et al., 2022). While six are prospective studies (Fung et al., 2024; Konieczynski et al., 2025; X. Li et al., 2025; Y. Li et al., 2025; Sandoval-Insausti, Blanco-Rojo, et al., 2020; Zhang et al., 2022), four are cross-sectional studies(Clayton-Chubb et al., 2024; Hallajzadeh et al., 2025; Hao et al., 2022; Zupo et al., 2023). For the assessment of ultra-processed food intake, eight papers used food frequency questionnaires (FFQ)(Clayton-Chubb et al., 2024; Fung et al., 2024; Hallajzadeh et al., 2025; Konieczynski et al., 2025; X. Li et al., 2025; Y. Li et al., 2025; Zhang et al., 2022; Zupo et al., 2023), one study used a 24-h dietary recall (Hao et al., 2022), and one study used a validated computerized face-to-face dietary history (DH-ENRICA) (Sandoval-Insausti, Blanco-Rojo, et al., 2020). For assessment of frailty, six papers utilized the Fried Frailty Phenotype (Fung et al., 2024; Hallajzadeh et al., 2025; Hao et al., 2022; Konieczynski et al., 2025; Sandoval-Insausti, Blanco-Rojo, et al., 2020; Zupo et al., 2023), one paper employed the 42-item accumulation deficit frailty index (X. Li et al., 2025), one paper used the APSirin in Reducing Events in the Elderly (ASPREE) deficit accumulation frailty index (Clayton-Chubb et al., 2024), and two papers used the handheld dynamometer (Y. Li et al., 2025; Zhang et al., 2022).

**Table 1.** Study characteristic.

| Studies | Country | Design | Sample size | Follow up (years) | Age : Mean (SD) | sex | Dietary assessments and exposure UPFs | Outcome measures | covariates |
| --- | --- | --- | --- | --- | --- | --- | --- | --- | --- |
| Sandoval-Insausti et al. (2020) | Spain | prospective | 1822 | 3.5 | 68.7 | 51.3% female | face-to-face dietary history (DH-ENRICA): UPF energy intake%; UPF gram per day/body weight (g/kg) | Fried frailty phenotype (Fried et al.,2003) | sex, age, education, marital status, tobacco consumption, and former-drinker status, chronic respiratory disease, coronary disease, stroke, osteoarthritis/arthritis, cancer, depression requiring treatment, and number of medications used. |
| Hao et al.(2022) | United states | Cross-sectional | 2329 | NA | 70.25(0.28) | 54.92% female | 24-h dietary recall: UPF energy intake; UPF energy intake%; UPF gram | Fried frailty phenotype (Fried et al.,2003) | age, sex, marital status, BMI, race, smoking, education level, poverty-income ratio, waist, and health conditions (diabetes, hypertension, cancer, etc.), and total energy intake |
| Zupo et al.(2023) | Italy | cross-sectional | 2185 | NA | 73.56(6.30) | 50.49% female | FFQ: UPF energy intake | Fried frailty phenotype (Fried et al.,2003) | age, sex, and education, IL-6, CRP, TNF-alpha, alcohol consumption, proteins/energy ratio, total daily energy intake, and multimorbidity |
| Li Y et al., (2025) | Singapore | prospective | 13570 | 21.2 | 53.21(6.1) | 58.9% female | FFQ: UPF weight proportion % | digital dynamometer: muscle weakness | age, sex, dialect group, educational level, alcohol frequency, smoking status, physical activity, sleep duration, total energy intake, caffeine, BMI, comorbidities, and diet quality |
| Konieczynski et al., (2025) | United states | prospective | 2547 | 10.8 | 60.3(8.9) | 55.1% | FFQ: UPF energy intake | Fried frailty phenotype (Fried et al., 2003) | baseline age, education, and sex, energy intake, multivitamin use, smoking status, self-rated health, history of diabetes, history of cancer and cardiovascular disease disorder, diet quality |
| Hallajzadeh et al., (2025) | Iran | cross-sectional | 368 | NA | 67.11(6.21) | 55.2% female | FFQ: UPF energy intake% | Fried frailty phenotype (Fried et al.2003) | socioeconomic status, sex, age, BMI, FAT, fat mass, AST, h-CRP, alkaline phosphatase, LDL-C, cholesterol, and triglycerides |
| Clayton et al.(2024) | Australia | cross-sectional | 12461 | NA | 76.9 | 54.4% female | FFQ: ASPREE-UPF score | The ASPREE deficit accumulation frailty index | age, sex, BMI, educational attainment, living situation, and current smoking and alcohol drinking status |
| Fung et al.(2024) | United states | prospective | 63743 | 26 | 60 | 100% female | FFQ: servings of UPF energy intake; UPF energy intake% | Fried frailty phenotype (Fried et al.2003) | age, energy intake, alcohol intake, baseline BMI, physical activity, smoking, postmenopausal hormone use, use of aspirin, diuretics, $\beta$ -blockers, calcium channel blockers, angiotensin-converting enzyme inhibitors, other antihypertensive medication; use of statins and other cholesterol-lowering drugs; use of insulin and oral hypoglycemic medication, SES, and diet quality |
| Zhang et al.(2022) | China | prospective | 5409 | >1 | 48.3 | 61.3% male | FFQ:<br>UPF energy intake% | handheld dynamometer:<br>low grip strength | baseline age, sex, BMI, smoking status, alcohol drinking status, education level, employment, and monthly household income, physical activity, family history of disease, depressive symptoms, hypertension, hyperlipidemia, diabetes, total energy intake, dietary supplement use, total protein intake, milk intake, healthy diet score, and baseline grip strength. |
| Li X et al., (2025) | Italy | prospective | 938 | 16.14 | 74(6.6) | 55.2% female | FFQ:<br>UPF consumption frequency | 42-item frailty index:<br>frailty baseline; frailty over time | age, sex, and years of education, BMI, smoking |

**Note:** FFQ = food frequency questionnaire, SD= standard deviation

### Quality of studies

According to NOS scores, we classified 8 studies(Fung et al., 2024; Hallajzadeh et al., 2025; Hao et al., 2022; Konieczynski et al., 2025; X. Li et al., 2025; Y. Li et al., 2025; Sandoval-Insausti, Jiménez-Onsurbe, et al., 2020; Zupo et al., 2023) as high quality, and 2 studies (Clayton-Chubb et al., 2024; Zhang et al., 2022) as fair quality. The details are provided in Supplementary Material **Table S1**.

### Narrative synthesis

Four cross-sectional studies reported mixed findings regarding the association between UPF intake and frailty. Hao et al. found that the energy (Kcal) of UPF intake was significantly associated with a higher risk of pre-frailty or frailty (OR = 1.04, 95% CI [1.01, 1.07], p=0.014), after adjusting for age, sex, BMI, race, total energy intake, diabetes, hypertension, and stroke (Hao et al., 2022). The energy proportion (%) of UPFs, as defined by the total energy of UPF intake/total energy intake during the day, was also associated with higher odds of pre-frailty or frailty (OR=1.06, 95% CI [1.01, 1.13], p=0.018). Although UPF intake expressed in grams was not associated with the combined effect of pre-frailty/frailty (OR=1.02, 95% CI [0.99, 1.04], p=0.144), it was significantly related to frailty alone (OR=1.04, 95% CI [1.00, 1.08]). In a study of 368 older adults, Hallajzadeh et al. found that UPF intake was significantly associated with frailty (OR = 2.15, 95% CI [1.13, 4.09], p = 0.019) (Hallajzadeh et al., 2025). Similarly, after adjusting demographic and lifestyle factors, Clayton-Chubb et al. found that a higher ASPREE-UPF score was associated with an increased odds of frailty (OR=1.10, 95% CI [1.06, 1.14], p<0.05), as assessed by the APSirin in Reducing Events in the Elderly (ASPREE) deficit accumulation frailty index (Clayton-Chubb et al., 2024). However, in an analysis of 2185 participants, Zupo et al. found that UPF intake was not significantly associated with nutritional frailty, defined as a combination of physical frailty and nutritional imbalance (OR = 1.26, 95% CI [0.86, 1.85], p > 0.05) (Zupo et al., 2023).

Six prospective studies showed a relatively consistent pattern, generally indicating that UPF intake was significantly associated with an increased risk of frailty or related outcomes. In a cohort of 1822 older adults followed for 3.5 years (Sandoval-Insausti, Jiménez-Onsurbe, et al., 2020), the energy percentage of UPFs was associated with an increased risk of frailty (OR = 3.67, 95% CI [2.00, 6.73], p < 0.001). When UPF was defined as grams per day/body weight (g/kg), a significant association with frailty was also observed (OR = 2.57, 95% CI [1.41, 4.70], p = 0.004). Furthermore, Fung et al. observed that an increase in UPF energy intake was associated with a 0.37-fold increase in frailty (HR=1.37, 95% CI [1.29, 1.46], p< 0.001) (Fung et al., 2024). When UPFs were defined as an energy percentage, there was also an increased risk of frailty in older adults (HR = 1.12, 95% CI [1.06, 1.18], p < 0.001). Similarly, in an Italian cohort of 938 participants (X. Li et al., 2025), higher UPF intake was significantly associated with both greater baseline frailty (β=0.026, 95% CI [0.01, 0.041], p=0.001), and steeper frailty progression over 16.1 years of follow-up (β=0.022, 95% CI [0.006, 0.027], p=0.006).

Consistent associations were also reported in Asian populations. Zhang et al. also found that UPF intake was associated with the risk of incident frailty in the Chinese population, defined as low grip strength (HR = 1.36, 95% CI [1.06, 1.74], p = 0.01) (Zhang et al., 2022). In the Singapore Chinese Health Study, midlife UPF intake was associated with an increased risk of muscle weakness in late life (OR=1.32, 95% CI [1.13, 1.53], p<0.001) (Y. Li et al., 2025). Among the six prospective studies, only one found no significant association between UPF intake and frailty over a 10.8-year follow-up in older adults (OR=1.01, 95% CI [0.95, 1.07], p=0.81) (Konieczynski et al., 2025); however, a significant association with grip strength was observed. All included studies adjusted for multiple covariates in their statistical models.

### Meta-analysis

The meta-analysis included six studies for which odds ratios (ORs) and 95% confidence intervals (Cis) were available. An additional four studies reporting HR, RR, or β were summarized qualitatively in the systematic review. Based on data from 22,821 participants, the pooled analysis showed that higher intake of ultra-processed foods was significantly associated with an increased risk of frailty (pooled OR = 1.43, 95% CI = [1.02-2.005], p = 0.041; Figure 2). We also observed significant between-study heterogeneity (Q(5) = 33.06, p < 0.001, I^2^ = 97.78%).

**Figure 2.**
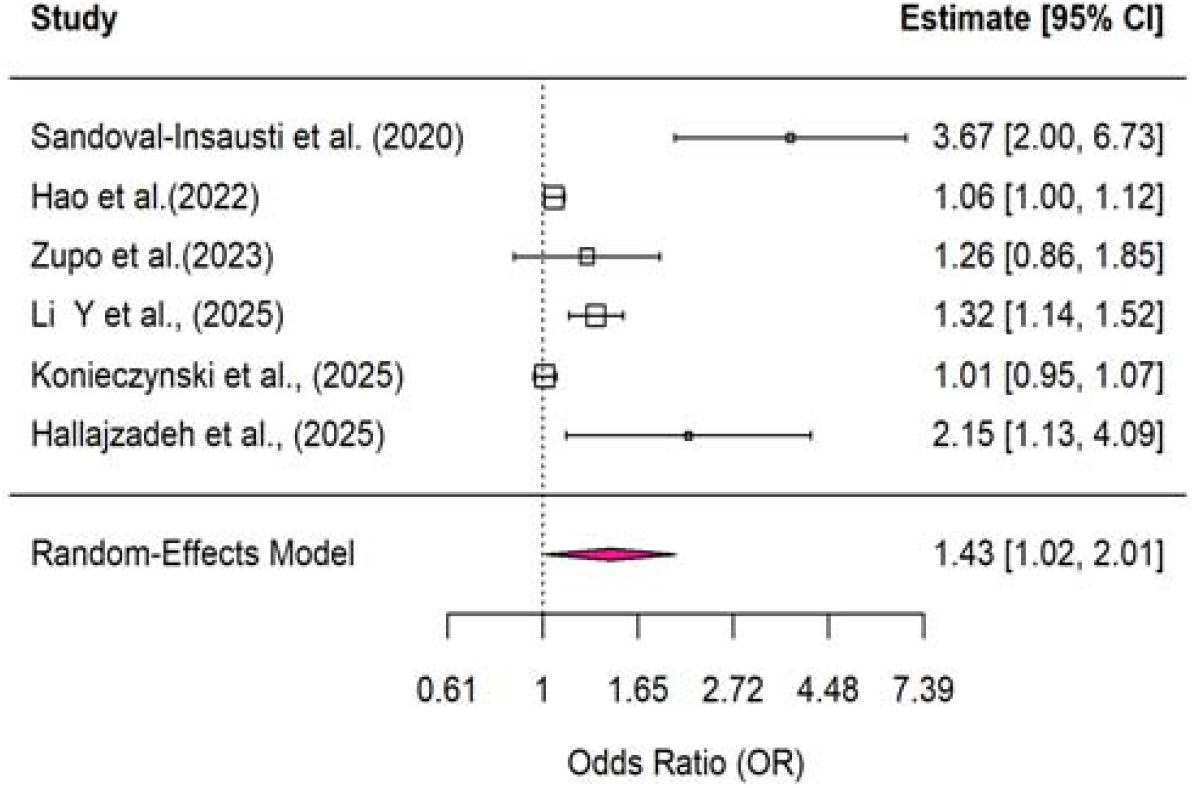
Forest plots from a random-effects model showing the association between high vs low ultra-processed food intake and risk of frailty. CI=confidence interval.

The funnel plot showed some degree of asymmetry (Supplementary **Figure S1**). Egger’s regression test indicated significant funnel plot asymmetry (z= 4.63, p < 0.001), suggesting potential small-study effects or publication bias. Furthermore, leave-one-out sensitivity analyses demonstrated that statistical significance was maintained in only a subset of models (Supplementary **Table S2**), indicating limited robustness of the pooled effect.

## Discussion

This study presents the first systematic review and meta-analysis examining the effect of ultra-processed foods on frailty risk. Our findings provide robust evidence that greater UPF intake is significantly associated with an increased prevalence of frailty among older adults. This suggests that interventions targeting UPF consumption may present as an important strategy for preventing age-related frailty and improving health outcomes.

Frailty has been consistently associated with adverse outcomes such as increased mortality, lower quality of life, dementia, and depression (Hoogendijk et al., 2019). Importantly, frailty is potentially preventable and dynamic rather than stable. Clinical factors such as cognitive impairment, lifestyle factors including poor diet, and biological factors such as chronic inflammation, have been identified as primary contributors to the onset and progression of frailty (Qin et al., 2023). Among those factors, diet is strongly associated with the pathophysiology of frailty. A meta-analysis of nine observational studies found that greater adherence to a healthy dietary pattern, such as high intake of vegetables, whole grains, and fruit, was associated with a reduced risk of frailty (Rashidi et al., 2019). Similarly, an analysis of 24996 adults from the UK Biobank found that a healthier plant-based diet was associated with a lower risk of frailty (Maroto-rodriguez et al., 2024). As observed across these studies, healthy, nutrient-dense diets are typically rich in phytochemicals with anti-inflammatory properties, which may confer protective effects against frailty (Lochlainn et al., 2021).

The consumption of UPFs is generally negatively associated with diet quality, suggesting that the association between UPFs and frailty may reflect the unhealthy nutritional profile of these foods. Supporting this review, a recent study reported a non-significant relationship between UPFs and frailty after adjusting for diet quality (Konieczynski et al., 2025). However, UPF intake remained significantly associated with annual changes in specific frailty components, including gait speed and grip strength. In contrast, a recent large-scale study of 63743 older adults in the United States found that, over a 26-year follow-up, 15187 participants developed frailty, and UPF consumption was significantly associated with frailty incidence even after adjusting for diet quality (Fung et al., 2024). These findings indicate that the detrimental effect of UPFs on frailty may remain independently of overall dietary quality. However, in the present review, only three included studies reported that diet quality was adjusted in the model, leaving its precise role unclear and highlighting the need for future research to disentangle these effects more rigorously.

One of the primary physiological mechanisms linking UPF consumption and frailty is chronic inflammation, which is particularly prevalent in older adults (Quetglas-Llabrés et al., 2023). The nutritional profile of UPFs is thought to promote chronic inflammation through excessive intake of rapidly absorbable carbohydrates, unhealthy fats, refined sugars, and salt, alongside inadequate consumption of fiber and protein, nutrients essential for maintaining metabolic and immune homeostasis (Ciaffi et al., 2025; Contreras-Rodriguez et al., 2023). A recent review revealed that greater UPF intake is associated with elevated inflammatory biomarkers, including C-reactive protein (CRP), interleukin-6 (IL-6), and Tumornekrosefaktor-alpha (TNF-α), potentially mediated by increased postprandial glycaemia, oxidative stress, and subsequent cytokine responses (Ciaffi et al., 2025; Teeman et al., 2016). Elevated inflammatory markers have been linked to age-related decline in psychological and physical functions (Pan & Ma, 2024). For instance, a longitudinal study found that IL-6 significantly predicted frailty one year later in prostate cancer patients (Buigues et al., 2020). Similarly, in healthy middle-aged and older adults, higher CRP levels significantly accelerated frailty progression over a 7-year follow-up period (Cheng et al., 2022). Furthermore, UPFs are typically produced with high levels of artificial additives, such as sweeteners and colorants, as well as processed substances that may disrupt immune homeostasis (Rondinella et al., 2025). Experimental evidence suggests that consumption of non-caloric sweeteners, such as sucralose, significantly alters inflammatory transcriptomic pathways in humans (Sylvetsky et al., 2020) and induces chronic inflammation in animal models (Bian et al., 2017). These findings collectively suggest that UPF consumption may promote frailty through inflammatory pathways.

Our findings have important implications for identifying modifiable targets for frailty prevention and treatment. Dietary interventions play an important role in preventing the progression of frailty (Hernández Morante, J. J., Gómez Martínez, C., & Morillas-Ruiz, 2019). In a randomized controlled trial, an intervention targeting good nutritional status and physical activity significantly reduced the risk of frailty by improving walking time among community-dwelling pre-frail older adults (Arradelles & Apiol, 2017). Another randomized controlled trial found that an adequate protein supplementation intervention significantly increased physical functioning by 5.9% compared with a control group among frail older adults (Randomized et al., 2013). Consistent with these dietary interventions, reducing UPF intake may be a promising strategy to mitigate frailty risk. While existing front-of-package label strategies primarily focus on nutrient content, studies examining labels that explicitly highlight levels of food processing remain limited. Notably, a recent study involving 600 adults in the United States found that exposure to an “ultraprocessed” label increased participants’ risk perceptions of UPFs and significantly reduced their intention to purchase these products compared with a control label (D’Angelo Campos et al., 2024). This suggests that processing-based labeling strategies may be effective in reducing UPF consumption and consequently frailty risk.

This study has several limitations. First, the relatively small number of included studies limited the possibility for subgroup analyses, such as distinguishing between cross-sectional associations and longitudinal effects of UPFs on frailty progression. Second, dietary assessment methods varied across studies. While eight studies employed a food frequency questionnaire, others used 24-h dietary recall or a validated computerized face-to-face dietary recall. This heterogeneity may have contributed to either over- or underestimation of effect sizes. Third, substantial between-study heterogeneity, attributable to study design and sample size, was observed. The findings should be interpreted cautiously.

In conclusion, this study summarized the available evidence on the relationship between ultra-processed foods and frailty risk and provided reliable evidence that UPFs significantly increase frailty progression. This study underscores the importance of public health strategies to reduce UPF consumption as a potential approach to preventing and reversing frailty in older adults.

## Data Availability

All data produced in the present study are available upon reasonable request to the authors

